# Clinical Impact of Metagenomic Next-Generation Sequencing of Plasma Cell-Free DNA for the Diagnosis of Infectious Diseases: A Multicenter Retrospective Cohort Study

**DOI:** 10.1101/19008755

**Authors:** Catherine A. Hogan, Shangxin Yang, Omai B. Garner, Daniel A. Green, Carlos A. Gomez, Jennifer Dien Bard, Benjamin A. Pinsky, Niaz Banaei

## Abstract

**Background:** Metagenomic next-generation sequencing (mNGS) of plasma cell-free DNA has emerged as an attractive diagnostic modality allowing broad-range pathogen detection, noninvasive sampling, and earlier diagnosis. However, little is known about its real-world clinical impact as used in routine practice.

**Methods:** We performed a retrospective cohort study of all patients for whom plasma mNGS (Karius test) was performed for all indications at 5 U.S. institutions over 1.5 years. Comprehensive chart review was performed, and standardized assessment of clinical impact of the mNGS based on the treating team’s interpretation of Karius results and patient management was established.

**Results:** A total of 82 Karius tests were evaluated, from 39 (47.6%) adults and 43 (52.4%) children and a total of 53 (64.6%) immunocompromised patients. Karius positivity rate was 50/82 (61.0%), with 24 (48.0%) showing two or more organisms (range, 2-8). The Karius test results led to positive impact in 6 (7.3%), negative impact in 3 (3.7%), no impact in 70 (85.4%), and was indeterminate in 3 (3.7%). Cases with positive Karius result and clinical impact involved bacteria and/or fungi but not DNA viruses or parasites. In 10 patients who underwent 16 additional repeated tests, only one was associated with clinical impact.

**Conclusions:** The real-world impact of the Karius test as currently used in routine clinical practice is limited. Further studies are needed to identify high-yield patient populations, define the complementary role of mNGS to conventional microbiological methods, and how best to integrate mNGS into current testing algorithms.

**Summary:** In a multicenter retrospective cohort study, we show that the real-world clinical impact of plasma metagenomic next-generation sequencing (mNGS) for the noninvasive diagnosis of infections is limited (positive impact 7.3%). Further studies are needed to optimize the impact of mNGS.

## Introduction

Metagenomic next-generation sequencing (mNGS) of pathogen nucleic acid in clinical samples has emerged as a promising one-test approach for hypothesis-free diagnosis of potentially all infectious etiologies. mNGS is currently orderable in select reference laboratories from plasma cell-free DNA (Karius, Redwood City, CA), from DNA and RNA in cerebrospinal fluid (CSF) (University of California, San Francisco) and DNA and RNA in respiratory secretions (IDbyDNA, San Francisco, CA) [1]. Of these, the Karius test has been commercially available the longest and is likely the most common mNGS send-out test in U.S. institutions. This assay is reported to detect and quantify pathogen cell-free DNA from 1,250 bacteria, DNA viruses, fungi and eukaryotic parasites [2], and has been employed for the diagnosis of invasive fungal disease [3, 4], community-acquired pneumonia [5] and infections in immunocompromised hosts [6, 7].

The availability of clinical metagenomic cell-free DNA sequencing has generated interest from clinical providers across disciplines given the noninvasive nature of testing, and the potential to pose an actionable diagnosis faster than through conventional microbiologic methods. However, little is known about the clinical impact of mNGS testing when used for routine patient care. A study assessing the clinical impact of mNGS from CSF in patients with suspected meningitis and encephalitis showed positive clinical impact in 8/204 (3.9%) patients tested [8]. In the present multicenter study, our aim was to comprehensively examine the real-world clinical impact of the Karius test ordered for all indications for the diagnosis of infectious diseases.

## Methods

### Ethics

This study was approved by the Institutional Review Boards (IRB) at Stanford University and Columbia Medical Center (CUMC). IRB was waived for this project for University of California Los Angeles (UCLA), University of Utah (UT) and Children’s Hospital of Los Angeles (CHLA).

### Karius Test

The Karius laboratory started to offer commercial testing in December 2016. Plasma from whole blood collected in a K2-EDTA or BD Vacutainer PPT tube (Becton Dickinson, Franklin Lakes, NJ) and separated <6 hours after draw was sent either fresh (within 4 days of blood draw) or frozen. All testing was performed by Karius per routine testing protocol as previously described [2].

### Study Design

We performed a retrospective cohort study of patients for all consecutive plasma mNGS samples sent-out for Karius test from 5 U.S. institutions (CHLA, CUMC, Stanford Health Care, UCLA and UT) from August 1^st^ 2017 to May 31^st^ 2019. Comprehensive chart review was performed by each site investigator for respective patients, and clinical impact solely based on the treating team’s interpretation of Karius results and their subsequent management decisions was assessed according to predefined objective grading criteria (Table 1) as to whether Karius test result had a positive, negative, or no clinical impact, or if impact was indeterminate. This study intended to assess the real-world impact of Karius test and therefore only the treating team’s decisions and actions were considered for impact assessment, and no retrospective clinical consensus assessment of impact and adjudication by a panel of experts was performed. Given that Karius testing may be reordered in the same patient for different reasons including patient monitoring, the main analysis was performed on the first Karius test per patient. A separate analysis was performed on repeat tests.

**Table 1.** Standardized criteria to assess clinical impact of the Karius test per the treating team.

| Category of clinical impact | Change in management | Clinical impact type |
| --- | --- | --- |
| Positive | Yes | New diagnosis based on Karius result and not confirmed by conventional microbiological methods |
|  |  | Earlier diagnosis based on Karius result and later confirmed by conventional microbiological methods |
|  |  | Karius result enabled avoidance of invasive surgical biopsy |
|  |  | Karius result enabled initiation of appropriate therapy |
|  |  | Karius result enabled de-escalation of therapy |
|  |  | Karius result enabled escalation of therapy |
|  | No | Karius result confirmed clinical diagnosis |
| Negative | Yes | Karius result led to unnecessary treatment |
|  |  | Karius result led to additional unnecessary diagnostic investigations |
|  |  | Karius result led to longer length-of-stay |
| None | No | Karius result showed new organism but result not acted upon |
|  |  | Karius result confirmed conventional microbiological diagnosis and not acted upon |
|  |  | Patient died before Karius result available |
| Indeterminate | Yes | Could not determine clinical impact from chart review |
|  | No |  |
|  | Indeterminate |  |

### Statistical Analysis

Statistical analysis was performed with Stata 15 (Stata Corp, TX, USA). Due to the descriptive nature of the review, a formal sample size calculation was not performed.

## Results

A total of 82 Karius tests were performed in unique patients cared for at UCLA (*n*=50 tests), SHC (*n*=21), CUMC (*n*=5), UT (*n*=3) and CHLA (*n*=3) (Table 2). There were 39 (47.6%) adults and 43 (52.4%) children, 52 (63.4%) males and 53 (64.6%) immunocompromised patients, most commonly solid organ transplant (17.1%) and primary immunodeficiency (13.4%) (Table 2). The mean turn-around-time from sample collection to result was 3.1 days (SD: 1.2), and the most common ordering providers were infectious disease physicians (74.4%), followed by oncologists (11.0%). The main indication for testing was fever of unknown origin (23.2%), suspected respiratory infection (13.4%), sepsis (9.8%), suspected endocarditis (8.5%) and febrile neutropenia (7.3%). The Karius positivity rate was 50/82 (61.0%), with 26 (52.0%) of these tests resulted as monomicrobial (18 bacterial, 5 viral and 3 fungal), and 24 (48.0%) showing 2 or more organisms (range, 2-8). There was no parasite detected in this cohort.

**Table 2.** Demographic and clinical characteristics of patients included in this study.

|  | **CUMC** | **SHC** | **UCLA** | **UT** | **CHLA** | **Total (%)** |
| --- | --- | --- | --- | --- | --- | --- |
| **Number of patients** | 5 | 21 | 50 | 3 | 3 | 82 |
| **Number of tests** | 6 | 22 | 64 | 3 | 3 | 98 |
| **Mean Age (SD)** | 12.0 (5.8) | 36.2 (24.3) | 20.5 (20.0) | 70.7 (8.4) | 4 (0) | 25.2 (23.3) |
| **Male sex (%)** | 2 (40.0) | 15 (71.4) | 29 (58.0) | 3 (100) | 3 (100) | 52 (63.4) |
| **Immunosuppression (%)** | 5 (100) | 14 (66.7) | 30 (60.0) | 3 (100) | 1 (33.3) | 53 (64.6) |
| Bone marrow transplant (%) | 4 (80.0) | 1 (4.8) | 2 (4.0) | 1 (33.3) | 0 | 8 (9.8) |
| Solid organ transplant (%) | 0 | 3 (14.3) | 11 (22.0) | 0 | 0 | 14 (17.1) |
| Active malignancy (%) | 1 (20.0) | 3 (14.3) | 4 (8.0) | 2 (66.7) | 0 | 10 (12.2) |
| HIV-1 (%) | 0 | 1 (4.8) | - | 0 | 0 | 1 (1.2) |
| Primary immunodeficiency (%) | 0 | 3 (14.3) | 7 (14.0) | 0 | 1 (33.3) | 11 (13.4) |
| Other (%) | 0 | 3 (14.3) | 6 (12.0) | 0 | 0 | 9 (11.0) |
| **Mean turnaround time**<br>**in days (SD)** | 2.8 (0.8) | 3.3 (1.9) | 3.0 (0.9) | 4.3 (0.6) | 2.3 (0.6) | 3.1 (1.2) |
| **Test indication for first test**<br>**per patient (%)** | - | - | - | - | - | - |
| Diagnosis | 5 (100) | 19 (90.5) | 46 (92.0) | 3 (100) | 3 (100) | 76 (92.7) |
| Rule-out infection | 0 | 1 (4.8) | 0 | 0 | 0 | 1 (1.2) |
| Monitoring | 0 | 1 (4.8) | 1 (2.0) | 0 | 0 | 2 (2.4) |
| Unknown | 0 | 0 | 3 (6.0) | 0 | 0 | 3 (3.7) |
| **Positive test results for first test per patient (%)** | 5 (100) | 12 (57.1) | 28 (56.0) | 3 (100) | 2 (66.7) | 50 (61.0) |

CHLA: Children's Hospital of Los Angeles; CUMC: Columbia University Medical Center; HIV-1: human immunodeficiency virus type 1; SD: Standard deviation; SHC: Stanford Health Care; UCLA: University of California, Los Angeles; UT: University of Utah.

The Karius test results led to positive or negative clinical impact in 9 (11.0%) tests. The impact was positive in 6 (7.3%) and negative in 3 (3.7%) (Table 3). Of these 9 Karius tests, 8 had organisms detected while 1 had no organism detected. Impact was none in 70 (85.4%) tests and indeterminate in 3 (3.7%). All 8 cases with positive Karius result and clinical impact involved bacteria and/or fungi; determination of the presence or absence of DNA viruses by plasma mNGS demonstrated no clinical impact in this study. Positive clinical impact was categorized as de-escalation of therapy (*n*=1), earlier diagnosis than conventional methods and initiation of appropriate therapy (*n*=2), new diagnosis not made by conventional methods and escalation of therapy (*n*=1), and new diagnosis and de-escalation (*n*=2). The 3 new diagnoses made by Karius test included presumptive diagnosis of an abdominal aorta mycotic aneurysm, a diagnosis of *Rhizopus oryzae* confirming disseminated mucormycosis, and identification of mixed oral flora in a case of blood culture-negative endocarditis, facilitating de-escalation of therapy. The 3 cases associated with negative clinical impact resulted in additional, unnecessary diagnostic investigations (*n*=2), and unnecessary treatment (*n*=1).

**Table 3.** Clinical data for cases with clinical impact after Karius test result.

| Study ID | Age/sex | Comorbidities | Clinical syndrome | Karius test result (molecules per microliter) | Clinical impact per treating team | Clinical impact type | Detail |
| --- | --- | --- | --- | --- | --- | --- | --- |
| SHC-11 | 57M | HIV/AIDS, CAD, CKD | Suspected mycotic aneurysm | *Enterococcus faecalis* (344)<br>*Prevotella melaninogenica* (176) | Positive | New diagnosis and de-escalation of therapy | Karius test result was used to de-escalate chronic suppressive therapy to amoxicillin-clavulanic acid, after a 6-week course of empiric vancomycin and ceftriaxone. Nine months later, after multiple clinical recurrences, vertebral tissue biopsy grew *Mycobacterium avium* complex. |
| SHC-13 | 47F | s/p combined heart and kidney transplant (2018) for anthracycline-induced cardiomyopathy, history of ALL | Acute flaccid paralysis (AFP) | *Aspergillus fumigatus* (qualitative result) | None/<br>Positive | None for AFP. Earlier diagnosis and initiation of therapy for fungal infection | Karius test identified *Aspergillus fumigatus* earlier than conventional methods and prompted chest imaging and respiratory cultures that confirmed invasive pulmonary aspergillosis. Serum galactomannan had been elevated (>2.00) in the week prior to Karius test. |
| UCLA-3 | 2M | Idiopathic acute liver failure, aplastic anemia, HLH | Recurrent fever | *Candida tropicalis*<br>(6,670) | Positive | Earlier diagnosis and initiation of therapy | Karius test result was used to initiate antifungal therapy with caspofungin 5 days earlier than the time of diagnosis by blood culture. |
| UCLA-20 | 10F | Liver/small bowel/colon/pancreas transplant (2018), TPN-dependent short gut syndrome | Jejunal perforation with necrotic fascia, subcutaneous abscess, perinephric fluid collection | *Rhizopus oryzae*<br>(169,935)<br>*Stenotrophomonas maltophilia*<br>(2,649)<br>*E. coli*<br>(234) | Positive | New diagnosis not made by conventional methods and escalation of therapy | Karius test diagnosed invasive mucormycosis, which was not made through conventional methods and which enabled targeted antifungal therapy. The patient was not treated for the bacterial organisms detected. |
| UCLA-47 | 6wk-old M | Newborn by emergency caesarean section, TAPVR s/p repair | Multi-organ dysfunction syndrome, coagulopathy, respiratory failure | Negative | Positive | De-escalation of therapy | Karius test result was negative and supported the clinical suspicion that patient's clinical decompensation was not infectious in etiology. This prompted discontinuation of empiric cefepime. |
| UT-2 | 75M | Metastatic carcinoma of unknown primary | Multifocal abscesses (lung, liver and kidney) | *Streptococcus parasanguinis*<br>(372)<br>*Morococcus cerebrosus*<br>(212)<br>*Neisseria sicca*<br>(96)<br>*Veillonella parvula*<br>(64)<br>*Prevotella melaninogenica*<br>(36)<br>*Propionibacterium acidifaciens*<br>(24)<br>EBV<br>(23) | Positive | New diagnosis not made by conventional methods and de-escalation of therapy | Karius test identified mixed oral flora, which was consistent with a periodontal source of infection.<br>This allowed narrowing of antibacterial therapy from vancomycin/piperacillin-tazobactam to amoxicillin-clavulanic acid for a total duration of 6 weeks of therapy. |
| CUMC-5 | 17F | GATA2-MDS s/p PBSC transplant (2018) | Fever, buccal lesions, hepatic lesions | *Trichosporon asahii*<br>(1,786)<br>*Enterococcus faecium* | Negative | Unnecessary treatment | Karius test result prompted additional antibacterial coverage that was not thought to be warranted per ID team. Patient |
|  |  |  |  | (1,567)<br>*Mucor indicus*<br>(1,199)<br>CMV<br>(763)<br>*Rhizopus delemar*<br>(107)<br>*Candida glabrata*<br>(75)<br>*Veillonella parvula*<br>(47) |  |  | was already known to have invasive mucormycosis. |
| SHC-8 | 39F | CVID | Chronic low-grade fever | *Prevotella bivia*<br>(23)<br>*Streptococcus pseudopneumoniae*<br>(23)<br>*Bacteroides fragilis*<br>(21) | Negative | Additional diagnostic investigations | Karius test result prompted an ED visit, and subsequent outpatient consultation with ID to outline management. The patient was not treated for the Karius test result. |
| SHC-14 | 20M | Refractory and recurrent marginal cell MALT-like lymphoma of ileum and colon | Investigation of lymphoma etiology | *Trichoderma atroviride*<br>(695) | Negative | Additional diagnostic investigations | Karius test result prompted concern by the treating team about a false positive result. Targeted fungal sequencing on |
|  |  |  |  | *Enterobacter*<br>*cloacae* complex<br>(96) |  |  | plasma was performed to try to<br>disprove result and was negative.<br>The patient was not treated for the<br>Karius test result. |

AIDS: acquired immune deficiency syndrome; ALL: acute lymphoblastic leukemia; AML: acute myeloid leukemia; AFP: acute flaccid paralysis; CAD: coronary artery disease; CHLA:
Children's Hospital of Los Angeles; CKD: chronic kidney disease; CUMC: Columbia University Medical Center; CVID: common variable immune deficiency; ED: emergency
department; F: female; GATA-2: GATA-2 gene; HIV: human immunodeficiency virus; HLH: hemophagocytic lymphohistiocytosis; ID: infectious diseases; M: male; MALT: mucosa-
associated lymphoid tissue; MDS: myelodysplastic syndrome; PBSC: peripheral blood stem cell; SHC: Stanford Health Care; s/p: status post; TAPVR: total anomalous pulmonary
venous return; TPN: total parenteral nutrition; UCLA: University of California, Los Angeles; UT: University of Utah; wk: week

A total of 25/82 (30.5%) tests underwent Karius testing with a pre-established microbiological diagnosis, with relative proportions varying by institution from 0 to 40%. The most common conventional diagnostic method establishing a microbiological diagnosis in this group was blood culture (*n*=9), tissue bacterial culture (*n*=6) and viral blood-based PCR (*n*=4). Of the 25 patients with a positive conventional diagnosis, 16 (64.0%) showed the same organism detected by the Karius test, with 7 of these showing other organisms in addition to the one that was reproducibly detected. The remaining 9 cases where Karius did not reproduce the result from conventional microbiological testing included bacteria (*Streptococcus viridans* (*n*=1), *Mycobacterium haemophilum* (*n*=1), *Mycobacterium chelonae* (*n*=1), *Mycobacterium bovis* (*n*=1)), DNA virus (adenovirus (*n*=1)), and fungi (*Candida parapsilosis* (*n*=1), *Candida lusitaniae* (*n*=1), *Coccidioides immitis*/*posadasii* (*n*=1), Mucorales (*n*=1)).

A subset of 10 patients underwent repeat testing, for a total of 16 additional repeated tests from UCLA (*n*=14), SHC (*n*=1) and CUMC (*n*=1). Review of the 16 repeat tests revealed 1 case associated with a positive clinical impact, where the repeat Karius test was performed 3 months after the initial one. This 2-year-old boy with a history of heart transplant was tested by Karius test shortly after an episode of *Stenotrophomonas maltophilia* bacteremia and positive respiratory cultures for the same organism. Karius testing confirmed *Stenotrophomonas maltophilia* and did not identify any other organism which prompted the team to de-escalate therapy.

## Discussion

Metagenomic next-generation sequencing has stimulated much interest among providers across medical subspecialties given its potential for broad-range pathogen detection. Furthermore, the noninvasive nature of plasma samples makes mNGS an attractive diagnostic modality for deep-seated infections that currently require invasive biopsy. However, the value of mNGS testing as a diagnostic tool is poorly understood. In this multicenter retrospective cohort study, we show that, as currently used in clinical practice, the real-world clinical impact of the Karius test is limited given that it resulted in no or negative impact in the majority (92.7%) of patients. This finding was consistent with that reported for mNGS on CSF where 196 of 204 (96.1%) tests did not have a clinical impact [8]. As it currently stands, in most academic and private health systems, microbiology and virology diagnostic laboratories offer a comprehensive menu of culture-based and molecular tests such that only few, typically complex, cases with infectious etiology will be left without a diagnosis. Important questions to consider now are whether mNGS has the diagnostic accuracy required to complement currently-available diagnostics, and how best to integrate mNGS into current testing algorithms. Published studies indicate CSF and plasma mNGS assays lack sensitivity compared to conventional methods and therefore cannot be used as stand-alone or rule-out tests [2, 8]. Thus, options include reserving mNGS as last resort for cases for which conventional microbiological testing has failed to provide an answer, or in some cases to consider more proximal testing in parallel to conventional tests. The decision to include mNGS testing should take into account the differential diagnosis, as well as the availability and accuracy of conventional methods. The specific example of lack of clinical impact of positive Karius results for DNA viruses in this cohort provides a compelling case for routine highly-available, highly-accurate molecular virology assays providing sufficient and actionable diagnostic information prior to Karius testing.

Plasma cell-free DNA may originate from the site of infection or colonization. Although Karius reports quantitative units, there is no threshold that distinguishes colonization from infection, and interpretation of results may therefore be clinically challenging, especially in immunocompromised hosts and patients with disrupted intestinal barrier. Clinical validation data recently reported on the Karius test from patients presenting to the emergency department at a tertiary hospital with suspected sepsis have demonstrated a positive percent agreement of 84.8% and negative percent agreement of 48.2% compared to all microbiological reference testing [2]. The finding that only 6 of 65 (9.2%) positive test results were positively impactful in the current study is consistent with an assay specificity that is insufficient to enable clinical impact when organisms are detected under routine practice. Even in certain cases where care was tailored to the positive Karius result, it is unclear if the reported organisms were the underlying drivers of pathology. For example, the patient with abdominal aorta mycotic aneurysm (Table 3; Study ID SHC-11) in whom antibiotic therapy was de-escalated to chronic suppressive therapy with amoxicillin-clavulanic acid based on a positive Karius result with *Enterococcus faecalis* and *Prevotella melaninogenica* presented with multiple clinical recurrences of back pain and eventually grew *Mycobacterium avium* complex (MAC) from a vertebral tissue biopsy 9 months later.

Patient selection practices may also have contributed to low clinical impact as almost one-third (ranging from 0 to 40% by institution) of patients had already received a microbiological diagnosis prior to testing. The only case in this cohort to have demonstrated clinical impact in the setting of a pre-established microbiological diagnosis is that of the repeat Karius test identifying *Stenotrophomonas maltophilia* only in a patient with known bacteremia with the same organism, leading to de-escalation of therapy. This highlights the important role of diagnostic stewardship in optimizing testing algorithms and preventing unnecessary testing. In situations where positive conventional test results precede the mNGS result, patient management (i.e., escalation or de-escalation of therapy) should be considered based on conventional microbiological diagnostics rather than waiting for confirmatory mNGS result.

The finding that 6/82 (7.3%) of Karius test results in this study led to positive clinical impact is in contrast to that recently reported in a single pediatric center in Chicago where 56/100 (56.0%) test results were considered clinically actionable [9]. This discrepancy may have been driven by differences in patient selection, in addition to differences in outcome assessment methodology. For instance, in our study, clinical impact was solely based on the treating team’s interpretation of Karius results and their subsequent management decisions; in contrast, in the above study, clinical adjudication was performed where ‘clinical relevance’ was unclear from the medical record. Inherent inter-provider interpretation differences of whether Karius results represent a clinically significant pathogen or colonization also likely contributed to findings. Other differences between studies included variability in testing indication, proportion of ordering infectious diseases providers, and proportion of patients with a known microbiological diagnosis pre-Karius testing. The low clinical impact proportion in the present study was closer to that reported for mNGS on CSF where 8/204 (3.9%) of tests led to positive clinical impact [8].

This study was strengthened by its multicenter design including 5 adult and pediatric sites, and by its assessment of real-world practice in presenting data from provider-initiated testing. However, there are limitations. Firstly, although we made significant effort to standardize the definition and application of clinical impact criteria, some of the cases included were very complex medically and interpretation of clinical impact may have been challenging to ascertain. The definition of impact was based on how providing physicians interpreted and reacted to test results. This may have over or underestimated the real impact of the Karius test result. Further studies with more comprehensive and objective measures of impact (for example, including antibiotic use, hospital length-of-stay, laboratory testing utilization) are needed to accurately quantify clinical impact. Secondly, approval criteria for send-out testing varied by institution and time period such that patient populations may have varied in their pre-test probability which may have influenced the assessment of clinical impact. Finally, given this was a convenience sample, not all patient populations are represented equally. For example, only one HIV-1-positive individual was included, which limits generalizability to this patient population.

In conclusion, plasma mNGS represents an attractive diagnostic modality due to its noninvasive nature and potential to provide an early and actionable diagnosis. However, this study has shown that the real-world impact of the Karius test as currently used in clinical practice is limited given that this testing resulted in no or negative impact in the majority (92.7%) of patients. Further studies are needed to better understand which patient populations may potentially benefit, define the complementary role of mNGS to conventional microbiological methods, and improve diagnostic interpretation algorithms. Furthermore, institutions should adopt an active diagnostic stewardship role to ensure optimal use of mNGS.

## Data Availability

The data have not yet been made available.

## References

1. Chiu CY, Miller SA. Clinical metagenomics. Nat Rev Genet 2019; 20(6): 341–55.

2. Blauwkamp TA, Thair S, Rosen MJ, et al. Analytical and clinical validation of a microbial cell-free DNA sequencing test for infectious disease. Nat Microbiol 2019; 4(4): 663–74.

3. Hong DK, Blauwkamp TA, Kertesz M, Bercovici S, Truong C, Banaei N. Liquid biopsy for infectious diseases: sequencing of cell-free plasma to detect pathogen DNA in patients with invasive fungal disease. Diagn Microbiol Infect Dis 2018; 92(3): 210–3.

4. Armstrong AE, Rossoff J, Hollemon D, Hong DK, Muller WJ, Chaudhury S. Cell-free DNA next-generation sequencing successfully detects infectious pathogens in pediatric oncology and hematopoietic stem cell transplant patients at risk for invasive fungal disease. Pediatr Blood Cancer 2019; 66(7): e27734.

5. Farnaes L, Wilke J, Ryan Loker K, et al. Community-acquired pneumonia in children: cell-free plasma sequencing for diagnosis and management. Diagn Microbiol Infect Dis 2019; 94(2): 188–91.

6. Fung M, Zompi S, Seng H, et al. Plasma Cell-Free DNA Next-Generation Sequencing to Diagnose and Monitor Infections in Allogeneic Hematopoietic Stem Cell Transplant Patients. Open Forum Infect Dis 2018; 5(12): ofy301.

7. Zhou Y, Hemmige V, Dalai SC, Hong DK, Muldrew K, Mohajer MA. Utility of Whole-Genome Next-Generation Sequencing of Plasma in Identifying Opportunistic Infections in HIV/AIDS. The Open AIDS Journal 2019; 13(1): 7–11.

8. Wilson MR, Sample HA, Zorn KC, et al. Clinical Metagenomic Sequencing for Diagnosis of Meningitis and Encephalitis. N Engl J Med 2019; 380(24): 2327–40.

9. Rossoff J, Chaudhury S, Soneji M, et al. Noninvasive Diagnosis of Infection Using Plasma Next-Generation Sequencing: A Single-Center Experience. Open Forum Infect Dis 2019; 6(8).

